# Aboriginal and Torres Strait Islander adolescent and parent perspectives on human papillomavirus vaccination through the School Immunisation Program in Queensland, Australia: a qualitative study

**DOI:** 10.1101/2025.10.08.25337639

**Authors:** Tamara L Butler, Kate Anderson, Ami Morseu-Diop, Julia M L Brotherton, Joan Cunningham, Allison Jaure, Gail Garvey, Evan AhWing, Vanessa Clements, Sonya Egert, Frances Lomas, Casey Ross, Lisa J Whop

## Abstract

Aboriginal and Torres Strait Islander peoples experience a greater burden of cervical cancer than other Australians. Human papillomavirus (HPV) vaccination can prevent cervical cancer, but uptake remains inequitable. We aimed to understand factors influencing Queensland Aboriginal and Torres Strait Islander adolescents’ participation in the school-based HPV vaccination program.

Using an Indigenist research approach and guided by a socio-ecological model for health promotion, Aboriginal and Torres Strait Islander voices were centred. In 2020-2021, we conducted Yarns (a culturally appropriate qualitative method) with 65 Aboriginal and Torres Strait Islander adolescents and 20 parents from 10 diverse Queensland schools. Yarns addressed understandings of HPV and HPV vaccination, resources, vaccine consent, vaccination experiences, and suggestions for improvement. Thematic analysis was conducted. Themes related to: HPV vaccination research during the COVID-19 pandemic; factors leading up to the vaccination clinic; and factors on the day of the vaccination clinic. The HPV vaccine was perceived as more trustworthy than the then-new COVID-19 vaccines. Despite varying knowledge of HPV and HPV vaccination, most adolescents and parents supported vaccinations to prevent diseases and benefit their health. Participants reported difficulties completing and returning the consent form and wanted more accessible information and communication, delivered in culturally appropriate ways. Adolescents reported needing privacy during vaccination, ways to manage heightened emotions triggered by the vaccination clinic, and the presence of trusted support people.

We provide recommendations to support Aboriginal and Torres Strait Islander adolescents to be vaccinated against HPV, such as flexible consent processes with assistance from Aboriginal and Torres Strait Islander support staff; availability of culturally safe educational materials; the presence of Aboriginal and Torres Strait Islander support staff at clinics, and optimal clinic set-up and scheduling. These recommendations can guide programmatic changes to increase HPV vaccination rates among Aboriginal and Torres Strait Islander adolescents, advancing equity in cervical cancer outcomes.

## Introduction

Cervical cancer is highly preventable and Australia is on the brink of eliminating this cancer as a public health problem (incidence <4 per 100,000) within the next decade.(1) Despite this, there is still significant within-country inequity, as Aboriginal and Torres Strait Islander peoples experience a disproportionately greater burden of cervical cancer than other Australians. Aboriginal and Torres Strait Islander peoples are the Indigenous peoples of Australia, making up 3.8% of population.(2) Incidence is 5.6 vs 12.3 per 100,000 females (2015–2019) and mortality is 1.3 vs 4.7 per 100,000 females (2017–2021) for Australian and Aboriginal and Torres Strait Islander populations, respectively.(3) A 68% reduction in cervical cancer incidence in Aboriginal and Torres Strait Islander women in Australia is required to reach the elimination threshold.(3) Human papillomavirus (HPV) causes almost all cancers of the cervix, a significant proportion of anal, vaginal, vulvar, penile and oropharyngeal cancers as well as genital warts.(4) HPV vaccination is a critical primary prevention strategy for these diseases and has been shown to prevent invasive cervical cancer.(5)

In Australia, HPV vaccination is recommended and funded by the National Immunisation Program (NIP) (6) and is provided routinely through school-based immunisation programs in each State and Territory, with catch up available through primary care. From 2007 it was offered to girls aged 12-13 years old, with a time-limited catch-up program for girls 14-26 years old, in a three-dose schedule using a quadrivalent (protecting against HPV types 6, 11, 16, and 18) vaccine. Boys were added to the NIP in 2013. In 2018 a two-dose nonavalent vaccine (protecting against HPV types 6, 11, 16, 18, 31, 33, 45, 52 and 58) was introduced.(7) Following the recommendation of the Australian Technical Advisory Group on Immunisation (ATAGI)(8), HPV vaccination has been delivered in a one-dose schedule using the nonavalent vaccine to all adolescents aged 12-13 years (Year 7)(6) since February 2023. It is typically administered via school-based immmunisation programs at the same time as the diphtheria, tetanus and pertussis vaccine and vaccination is recorded in the Australian Immunisation Register.

The National Strategy for the Elimination of Cervical Cancer in Australia sets targets across three pillars (HPV vaccination, cervical screening, and treatment) to achieve cervical cancer elimination by 2035. The HPV vaccination target is to vaccinate 90% of Australian girls and boys (9). Aboriginal and Torres Strait Islander people are one of the priority populations identified in this Strategy. There are specific priorities and actions relating to the need for tailored solutions to support the “acceptability, community support for, experience and understanding of school-based HPV vaccination among Aboriginal and Torres Strait Islander adolescents.” (pg 29,9). National data indicate that in 2023, 80.9% and 75% of Aboriginal and Torres Strait Islander girls and boys, respectively, had HPV vaccination coverage by age 15 years compared to 84.2% and 81.8% of all Australian girls and boys, respectively.(10)

Aboriginal and Torres Strait Islander people residing in the state of Queensland, located in the north-east of Australia, are the focus of the current study. About 4.6% of the Queensland population identify as Aboriginal and/or Torres Strait Islander. Queensland is lagging behind the nation in HPV vaccine coverage by 15 years of age, with 79.5% and 74.2% coverage of at least one dose amongst Aboriginal and Torres Strait Islander girls and boys, respectively, and 81.7% and 79% coverage of all Australian girls and boys, respectively, in 2023.(10) These state-level figures likely mask significant variability in vaccination rates in Queensland schools. Data from other Australian jurisdictions indicate that lower levels of HPV vaccination initiation occurred in schools that: are small, in remote locations, in lower socioeconomic areas, have lower attendance rates, and/or have a higher proportion of Aboriginal and Torres Strait Islander students and students with a disability (11). Furthermore, HPV vaccination coverage has continued to drop following the COVID-19 pandemic,(3, 10) undoing modest but steady increases in pre-pandemic HPV vaccination coverage.(3) Together this evidence indicates an urgent need to increase HPV vaccination uptake among all populations, in particular Aboriginal and Torres Strait Islander adolescents, to meet the 90% target set by the National Strategy.

Some of the barriers to Indigenous peoples’ uptake of HPV vaccination globally, include: distrust and avoidance of government organisations and pharmaceutical companies as a colonial legacy of maltreatment; need for greater understanding, accessibility and availability of culturally appropriate information and education about HPV, the vaccine and its benefits; mistrust in the vaccine and concerns about vaccine safety; cultural taboos and beliefs about the vaccine promoting sexual behaviours; and colonial systems and processes such as complex informed consent procedures.(12–15) Reported enablers for Indigenous peoples’ uptake of the vaccine globally, include: systems, processes, and programs with a specific focus on equity; families prioritising preventative health for their children; family experiences of cancer; multigenerational support and involvement in discussion and promotion of the vaccine and the collective involvement of the community in the decision making and health promotion; greater individual and community level knowledge of HPV and HPV vaccine; recommendations of trusted health organisations and health care professionals; and government subsidised vaccine programs, among other enablers. (13–15)

A systematic review of barriers and supports for uptake of HPV vaccination among Indigenous peoples globally found no qualitative studies reporting the perspectives of both Aboriginal and Torres Strait Islander adolescents and/or parents on Australian school based HPV vaccination programs. (15) Furthermore, there was a concerning lack of research that involved or consulted with Indigenous communities in their research approach.(15) As Australia moves toward cervical cancer elimination, it is critical that culturally appropriate and Aboriginal and Torres Strait Islander-led research be conducted with Aboriginal and Torres Strait Islander peoples and communities to ensure elimination includes us. To do so, we must understand and explore Aboriginal and Torres Strait Islander peoples’ perceptions and experiences of the school-based HPV vaccination program and identify factors to improve uptake.

The current research fills this gap, reporting on the ‘*Yarning about HPV Vaccination’* project that was conducted for, with, and by Aboriginal and Torres Strait Islander people and communities in the state of Queensland.(16) This study reports the findings of qualitative research conducted with Aboriginal and Torres Strait Islander adolescents and parents. The aim of this study was to understand the factors that influence the participation of Aboriginal and Torres Strait Islander adolescents in the school-based HPV vaccination program.

## Methods

### Study approach

The approach is outlined in the protocol.(16) Briefly, the research design, data collection and analysis were guided by an Indigenist research approach(17, 18) and the socio-ecological model for health promotion.(19) These approaches ensured Aboriginal and Torres Strait Islander voices and priorities were centred in the leadership, conduct and benefit of the research and that the complex interplay of individual, interpersonal, institutional, community and policy factors that impact on HPV vaccination uptake and completion were considered. The research aims for the project were developed to address priorities identified by Aboriginal and Torres Strait Islander Chief Investigators(16) and Indigenous researchers globally to address the urgent need to eliminate cervical cancer among Indigenous peoples.(14)

Reporting adheres to the Consolidated criteria for reporting qualitative research (COREQ, see S1 File)(20) and the Consolidated criteria for strengthening reporting of health research involving Indigenous peoples (the CONSIDER statement, S2 File).(21)

### Reflexivity

The research team acknowledge the significant role of identity and reflexivity in conducting qualitative research and research with Aboriginal and Torres Strait Islander peoples. Together, the research team, including both Aboriginal and Torres Strait Islander and non-Indigenous researchers, shares a wealth of knowledge and experience in research relating to Aboriginal and Torres Strait Islander health, health equity, cervical cancer, and cancer prevention, screening, and vaccination.

TB is an Aboriginal woman (Undumbi) and early career researcher with a focus on addressing health disparities in cancer screening and prevention among Aboriginal and Torres Strait Islander peoples, particularly for gynaecological cancers. AMD is a Torres Strait Islander (Kemer Kemer Meriam, Kulkalgal) woman and research officer working in cervical cancer elimination among Aboriginal and Torres Strait Islander people. JMLB is a female non-Indigenous public health physician and Professor of Cancer Prevention Policy and Implementation with particular expertise in HPV vaccination and cervical screening. KA is a female non-Indigenous, Australian senior researcher experienced in conducting collaborative qualitative research with Aboriginal and Torres Strait Islander researchers and communities. JC is a non-Indigenous equity-focused social epidemiologist with extensive experience in research relating to cancer in Aboriginal and Torres Strait Islander people. AJ is a non-Indigenous woman who has experience in patient-centred outcomes research. GG is an Aboriginal (Kamilaroi) senior researcher and Professor of Indigenous Health Research. EAW is an Aboriginal man born on Kaladoon Country with connections to Garawa, Waanyi, and Alyawarre on his mother’s side and Kalkadoon, Mayi-Yapi and Kukatj peoples on his father’s side. He has a strong interest in supporting Aboriginal and Torres Strait Islander young people’s health and wellbeing. VC is an Aboriginal (Yidinji) woman with a background in epidemiology and health care provision managing the Clinical Engagement Team in Queensland Health First Nations Health Office. SE is a Noonuccal Goenpul woman, living and working on Yuggera Country in Inala, Queensland. She previously worked at the Southern Queensland Centre of Excellence in Aboriginal and Torres Strait Islander Primary Health Care and coordinated the Inala Community Jury. Sonya is now CEO at Inala Wangarra. FL is a Kamilaroi woman working as Women’s Program Director at Inala Wangarra with a strong interest in supporting community programs and engagement. CR is a Bindal woman and supports Aboriginal and Torres Strait Islander students and their families. LJW is a Wagadagam, Gumulgal woman and senior researcher and epidemiologist. She has significant expertise in cervical cancer prevention and Aboriginal and Torres Strait Islander health.

### Governance

An Aboriginal and Torres Strait Islander Steering Committee (EAW, VC, SE, FL, and CR) was formed to advise and monitor the *Yarning about HPV Vaccination* project to ensure it was conducted in an ethical and culturally safe manner. The Steering Committee provided interpretation and feedback on study findings. All members were Aboriginal and Torres Strait Islander people and the group included multidisciplinary expertise across lived experience, primary health care, cervical cancer and screening, education, youth support services, and community development programs. It included both male and female members. Due to disruptions to research caused by the pandemic, the first meeting was convened in early 2021, shortly after data collection commenced. The group met at significant milestones throughout the project, including during data collection, when data collection concluded, and to discuss preliminary findings. Steering Committee members were remunerated for their time.

### Recruitment and participants

In this article, we use “adolescents” to refer to Year 7 (12–13-year-old) Aboriginal and Torres Strait Islander students eligible for HPV vaccination and “parents” to inclusively refer to parents, caregivers, residential college guardians, and kin and family members (e.g., Aunties, Uncles or grandparents) who look after adolescents and have authority to provide consent for HPV vaccinations.

School recruitment was purposive and followed the eligibility criteria set out in the Protocol.(16) Briefly, the eligible schools were located in Queensland and had a relatively high proportion of Aboriginal and Torres Strait Islander student enrolments (identified using the Australian Curriculum, Assessment and Reporting Authority (ACARA)(22) School Profile data. School recruitment continued until 10 Queensland schools (see S3 File) had been recruited representing a maximally diverse range of student gender (female, male, mixed), school sector as per ACARA School Profile data ((22)Catholic, government, independent), and geographic remoteness (remote, regional, urban). As recruitment of a maximally diverse sample of 10 schools was prioritised, data collection continued after data saturation was achieved.

The School Principal provided written informed consent for their school’s participation in the study and nominated a school representative to liaise with the research team regarding participant recruitment and research logistics. Where possible the school representative was, or put the research team in contact with, a local Aboriginal and Torres Strait Islander staff member who could advise on local protocols and acceptable ways of conducting research. The school representative emailed or sent home hard copies of the study information and consent forms to Year 7 Aboriginal and Torres Strait Islander students and their parents. Informed consent for the study (collected via online form, signed hard copy, or over the phone audio-recorded formats depending on participant preference and convenience) was obtained from all participants. Both adolescent and parent consent was required for an adolescent to participate.

Both adolescents and their parents were invited to participate in the study: however, it was not a requirement that both the adolescent and their parent participate. Either adolescent and/or parent could participate.

As school representatives conducted the recruitment, we are unable to report how many people declined to participate in the study. No participant withdrew from the study after commencing.

At all schools, opportunities for reciprocity essential to research in Aboriginal and Torres Strait Islander communities and relationality between Aboriginal and Torres Strait Islander people were offered as part of the recruitment process. This involved offering services or resources that would be beneficial to Aboriginal and Torres Strait Islander students and aligned with the school’s priorities, and was up to the discretion of the school. For example, one school requested merchandise for a prize pool for school attendance, while another requested funding to support National Aborigines and Islanders Day Observance Committee (NAIDOC) week (an annual celebration of Aboriginal and Torres Strait Islander identity and culture) food and activities.

### Data collection

Data collection occurred between 15 October 2020 and 1 December 2021, inclusive. Wherever possible, research visits were timed to coincide with the school-immunisation team visit. At the time of the research, HPV vaccination was delivered in a two-dose schedule, with doses delivered 6 months apart. Researchers conducted observation of the clinic and collected field notes regarding topics such as the room layout, weather, significant events during the day (e.g., timing of other school activities), COVID-19 impacts, support people present, and observed barriers and facilitators to vaccination. This provided rich contextual information for Yarns with participants and data analysis.

Aboriginal and Torres Strait Islander researchers collected data, with experienced qualitative researchers TB (female, Aboriginal) and LJW (female, Torres Strait Islander) conducting most of the Yarns. In several communities, local Aboriginal and Torres Strait Islander researchers assisted in data collection: RF (male, Aboriginal), EA (male, Aboriginal) and ES (female, Torres Strait Islander). Qualitative research training was provided to community researchers, with further training and learning provided during field work. Community researchers were remunerated for all training, preparation, and field work. Local researchers advised the research team on appropriate protocol and practices. When requested by the school site, gender of the researcher was matched to the gender of the participant for Men’s and Women’s business.

Data was collected through semi-structured Yarns (individually) or Yarning Circles (in a group) with participants. Yarning is a culturally appropriate qualitative method providing Aboriginal and Torres Strait Islander participants with a safe and culturally strong space to share stories while respecting cultural protocols.(23) Using Social Yarning,(23) researchers first established relationships with participants by sharing each other’s Country, kin and family ties, and connecting with participants through culture. Food was shared.

Participants first completed a brief survey covering demographic and HPV vaccination information: Aboriginal and Torres Strait Islander identification, age, gender, residential location, religion, day or boarding study status, when the student had started attending the school, whether the adolescent had received any vaccinations in the previous year, whether the adolescent received any HPV vaccinations this year, number and location of each of the HPV vaccinations. Parents and adolescents completed the same survey questions with age-appropriate adjustments.

Research Yarns were then conducted using a semi-structured Yarning guide developed for this project (see S4 File). Yarns focused on several key topics: views and understanding of HPV and HPV vaccination, information, decision-making and consent, experience of the school vaccination clinic, impacts and understanding of COVID-19 and COVID-19 vaccination, and suggestions for improving HPV vaccination for Aboriginal and Torres Strait Islander adolescents. We adjusted the Yarning guide slightly after the first school visit to better explore COVID-19 impacts and vaccination.

We conducted Yarns with adolescents in-person at the school and Yarns with parents over the phone. All Yarns were audio-recorded with the consent of the participant; where consent was not provided, the researcher took notes. Yarns ranged from 5 to 46 minutes long. Generally, a school staff member was present to provide supervision for the Yarns with adolescents but maintained distance to give the participant privacy.

After each Yarn concluded, we provided age-appropriate information about HPV and HPV vaccination. All adolescents and parents received a gift voucher to the value of $30 or $50, respectively. Gift vouchers were tailored to the preferences of the school and community; for example, music streaming services, phone credit, the school canteen, department stores or grocery stores.

Yarns were transcribed verbatim and were not returned to participants for feedback or correction. Physical completed forms and surveys were stored in a locked filing cabinet and digital forms were stored on secure servers with access restricted to project team members.

### Data analysis

We employed thematic analysis using a constructivist approach.(24) The socio-ecological model for health promotion(19) was used as a framework to ensure individual, social, structural and environmental factors were considered in analysis. Importantly, Aboriginal and Torres Strait Islander researchers and the Steering Committee led the coding and interpretation of data ensuring appropriate cultural criticality and strengths-based lens was applied, aligning with an Indigenist research approach.(17, 18)

Aboriginal and Torres Strait Islander researchers TB, LJW, and AD created initial coding themes by reflecting on several transcripts, researcher experiences conducting Yarns, and with reference to the socio-ecological model for health promotion(19) levels of influence. TB and KA then piloted coding 4 transcripts. Following discussion and reorganisation of the coding structure with KA and LJW, TB coded the remaining transcripts and synthesised the findings, including data from observations of clinics and field notes. Preliminary findings were presented to the Aboriginal and Torres Strait Islander Steering Committee and their feedback was integrated. Transcripts and observational field notes were managed with NVivo Software.

### Ethics approval

Ethical approval for the study was provided by the Human Research Ethics Committee (HREC) of the NT Department of Health and Menzies School of Health Research (2019–3484), Aboriginal Health and Medical Research Council HREC (1646/20), Australian National University HREC (2020/478), Townsville HHS HREC (HREC/QTHS/73789), the University of Queensland HREC (#2021/HE002276), and Far North Queensland HREC (#HREC/2021/QCH/78996). All research adhered to the relevant ethical guidelines and regulations.

Approvals to conduct research were provided by Department of Education Queensland Research Services (550/27/2242), Edmund Rice Education Australia, Diocese of Toowoomba Catholic Education, Diocese of Townsville Catholic Education (2020-05).

All participants provided informed consent in either written or audio-recorded verbal format. Verbal consent, documented through audio recording, was required for interviews conducted over the phone. Adolescent participants required parent consent and personal consent to participate.

## Results

Participant characteristics are provided in Table 1. Most adolescents (median age = 12) had received at least one HPV vaccination during the school year, and almost all indicated that they received it at the school immunisation clinic. Most parents (median age = 43, range 32-69 years old) reported their adolescent had received at least one HPV vaccination dose.

**Table 1.** Participant characteristics.

|  | Number of<br>adolescents (%) | Number of Parents<br>(%) |
| --- | --- | --- |
| Total | 65 | 20 |
| **Indigenous Identification** |  |  |
| Aboriginal | 35 (54) | 6 (30) |
| Torres Strait Islander | 15 (23) | 3 (15) |
| Both Aboriginal and Torres Strait<br>Islander | 15 (23) | 3 (15) |
| Non-Indigenous | - | 8 (40) |
| **Gender** |  |  |
| Male | 25 (38.5) | 4 (20) |
| Female | 40 (61.5) | 16 (80) |
| **Remoteness** |  |  |
| Major cities | 7 (11) | 0 |
| Regional | 33 (51) | 10 (63) |
| Remote | 25 (38) | 6 (37) |
| **Religion** |  |  |
| No religion | 37 (57) | 7 (35) |
| Catholic | 12 (18) | 6 (30) |
| Anglican | 3 (5) | 4 (20) |
| Other religion | 6 (9) | 3 (15) |
| Don't know | 5 (7) | - |
| Missing data | 2 (3) | - |
| **Day/Boarding Student** |  |  |
| Day student | 51 (78) | 19 (95) |
| Boarding student | 14 (22) | 1 (5) |
| **Has adolescent received any HPV vaccinations this year?** |  |  |
| Yes | 51 (78) | 15 (75) |
| No | 12 (18) | 2 (10) |
| Don't know | 2 (3) | 1 (5) |
| Missing data | - | 2 (10) |
| **Number of HPV vaccinations (n=51 adolescents, n= 15 for parents)** |  |  |
| 1 | 39 (76) | 13 (87) |
| 2 | 10 (20) | 1 (7) |
| Missing data | 2 (4) | 1 (7) |

Themes identified were: 1) HPV vaccination research during the COVID-19 pandemic (with subthemes: disruptions to HPV vaccination clinics; parents trusted the HPV vaccine more than COVID-19 vaccine), 2) factors leading up to the HPV vaccination clinic (with subthemes: overarching support for vaccines; challenges with consent forms; parents led decision-making; need for accessible information and improved communication; HPV vaccination triggered ‘tricky’ conversations) and 3) factors on the day of the HPV vaccination clinic (with subthemes: emotions ran high; need for privacy; physical characteristics of the clinic; trusted support people; and interactions with immunisers were brief and impersonal). Supporting quotes are provided and marked with the participant type, participant identification number and school identification number.

### HPV vaccination research during the COVID-19 pandemic

#### Disruptions to HPV vaccination clinics

Field work for this study was scheduled to begin in March 2020 but was delayed by COVID-19 lockdowns and travel restrictions. When field work recommenced in late 2020, it was clear that the pandemic had influenced how adolescents and parents understood infectious diseases and the purpose of vaccinations. It also caused major disruptions to the delivery of school based immunisations. Over the course of data collection (2020 to 2021), participants demonstrated an increasing awareness and understanding of vaccines, side effects, and vaccine development. Fieldwork coincided with the rollout of COVID-19 vaccination in Australia with heightened public debate being broadcast on its efficacy and safety. Because of these events, many participants reflected on COVID-19 vaccinations in contrast with HPV vaccination and this influenced participants’ perceptions of the HPV vaccination program.

In some schools, boarding students had been sent home to families when lockdowns commenced in early 2020 and therefore missed out on the first dose of the HPV vaccine (scheduled in most schools early in the school year; in Australia the school year is from late January to December). They instead received their first dose when day student peers were receiving their second dose. In several schools that we visited toward the end of 2021, the COVID-19 vaccination rollout for children prevented some adolescents from receiving the HPV vaccination at school. This was mainly due to the clinical guidelines at the time preventing individuals from receiving the two vaccines close together. Additional catch-up HPV vaccination clinics were scheduled but were expected to be hampered by declining school attendance toward the end of the year. Some adolescents who received the HPV vaccination reported confusion about whether they had received the COVID-19 vaccine instead.

#### Parents trusted the HPV vaccine more than the COVID-19 vaccine

Parents widely viewed the HPV vaccine as trustworthy and safe because of its long history and track record of being administered in schools. Several parents made direct comparisons to the COVID-19 vaccine, with the HPV vaccine seen as routine and regular and therefore trustworthy.

> *[HPV] didn’t seem like anything out of the ordinary or there was something really new, like, for instance, COVID’s really new and the vaccination’s really new, so everyone was alarmed about whether or not they were going to get vaccinated. So, from memory, what I read on the form, it didn’t seem like it was talking about, like, a new type of disease or something that was not common for Indigenous people. So I kind of – there was no alarms*. Parent ID6688, S09.

### Leading up to HPV vaccination clinic

#### Overarching support for vaccines

Parents and adolescents held a range of knowledge about the benefits of the HPV vaccine. Some specifically cited benefits of the vaccine being able to prevent cancer, with many participants focusing on the benefits for girls specifically and to prevent cervical cancer. Few mentioned the broader benefits extending to boys and girls: a handful mentioned prevention of genital warts, and no participants mentioned prevention of other cancers caused by HPV. A few parents were aware of the sexually transmitted nature of HPV but very few had discussed this with their child.

Despite this, most adolescents and parents supported the idea of vaccinations to prevent diseases and benefit their health (with some reservations about the COVID-19 vaccination as explained above) and felt that being vaccinated would keep adolescents happy and healthy into their adult lives. Support for vaccines was driven by health benefits, trust, norms, and convenience.

> *Because it will stop me from getting a disease that I don’t want and that could possibly kill me. Adolescent, ID1656, S04.*

> *I just believe [vaccines] do more good than harm. And if they can help to stamp out some kind of sickness like polio or smallpox or what it has done in the past, that can only be a good thing.* Parent, ID4660, S02.

Some parents perceived their adolescent to be at greater risk of disease due to being an Aboriginal or Torres Strait Islander person, therefore making the vaccine doubly important.

> *Getting vaccination for your children is the best thing to do to keep them safe, keep them healthy, especially being Aboriginal. So I want to shield them against anything I possibly can.* Parent, ID8400, S05

Many parents conveyed a sense of trust in the school, medical experts, and the government health department which translated to support for the vaccine program.

> *…if the Health Department tells me my kids need a vaccine, I usually listen and go okay, it’s for the benefit of my kids to be healthier or prevent them from getting something.* Parent, ID3407, S03.

Parents and adolescents relied on norms in the school and community to form opinions about having the HPV vaccine. Knowing that “*everyone else*” (Parent, ID6688, S09) was getting the vaccine helped parents to make the decision to consent for the vaccine and adolescents also felt reassured that most of their classmates were having the vaccine.

Parents also relied on broader vaccination norms in their families to justify their decisions to vaccinate adolescents.

> *Well, I was vaxxed as a kid, my whole family was vaxxed. There’s never been any reason not to.* Parent, ID4660, S02

Some families also reported close family experiences of cancer that led them to support the vaccination.

Finally, many parents found the school program to be a convenient way for their child to receive vaccinations. It saved parents from taking leave from work or time out of a busy schedule and ensured their child did not miss school.

For adolescents, harms associated with the HPV vaccine largely related to worries about pain and discomfort. These were not specific HPV vaccination and instead related to all vaccines. Parents also had concerns about the possibility of side effects and adverse reactions.

> *…I was thinking, did I do the right thing letting my child and signing the form? Or was there an allergy my child could get to this vaccine? And now I’ve got a son that’s allergic to Penicillin, so the things there like that risk that you’re taking. … And the unknown of what could happen. I mean she could have a reaction, you don’t know.* Parent, Yarning Circle 1, S02.

#### Challenges with consent forms

In Queensland, parents provide consent for the School Immunisation Program via a hard copy Queensland Health consent form, which is provided to parents via schools alongside an information sheet about the vaccines for which the adolescent is eligible.(25) The completed consent form is generally returned to the school by the student and passed onto the relevant health authority organising the clinic.

Both adolescents and parents reported difficulties in getting the vaccination consent form home, completed, and returned to school, with forms frequently misplaced, lost, or forgotten. These difficulties were usually associated with adolescents forgetting, misplacing the forms and/or not returning them to school once signed. Boarding school adolescents had limited opportunity to discuss the vaccination with their parents. Parents sometimes forgot to sign forms. Some adolescents hinted at intentionally leaving the form in their bag or losing it to avoid it being completed.

To overcome these challenges, parents searched school bags or had a system for forms that needed to be signed to be placed in a particular location. At one school, staff members sometimes met parents at the school gate during drop-off or pick-up to ensure the form made it home.

Parents suggested that receiving the paperwork directly via mail or email may be more effective than relying on adolescents. Another suggested providing consent over the phone to someone that could be trusted. Another parent suggested that a phone call from a staff member of an existing Aboriginal and Torres Strait Islander program their child was involved with would help ensure forms were completed and returned.

> *I have three boys and trying to have them bring forms home is like pulling teeth. So if someone calls and says, hey, a form’s gone home for something quite important, have you got it, have you sent it back? I’d be like, yes, I’ve got it, it’s gone back on this day, or no, I haven’t got it, I’m just going to have to rummage through a school bag to find it.* Parent, ID8400, S05

#### Need for accessible information and improved communication

Some parents had a cursory memory and understanding of information they received about the vaccination, which vaccines they had consented for and their purpose. Busy family lives prevented some parents from reading the forms closely and feeling fully informed. This led to feelings of guilt.

> *I feel like a dodgy parent, but anyway … Oh, because I can’t – don’t know what vaccinations she actually did have. I know – remember thinking at the time, when I signed it, that she should get them. So, it would have been something that we had thought, yeah, we should definitely get this done, but I’m not really sure or I can’t recall exactly which ones it was for at the time they had.* Parent, ID5477, S01

Many that remembered seeing the government-issued standard information sheet described it as too dense, text heavy, and needing to be written in plain language because “we’re not all experts in the medical field.”(Parent, ID3407, S03). The amount of detail on the forms and busy family life was a barrier to some parents reading it.

> *… there was some information, a brochure on it. I obviously couldn’t care to read it such in detail with it, so I just signed it off …* Parent, Yarning Circle 1, S02

Adolescents commonly received information from teachers or nurse immunisers about the vaccine on the day of the clinic or just before the vaccine was administered. This timing made it difficult for adolescents to take in and retain the information due to the heightened emotions of the day. Adolescents also reported seeking out more information about the vaccine in an opportunistic manner from teachers.

> *At break, with me and my friend, because we saw [the science teacher], because he was on duty. And then she said that I had to get my needle, and then he said, “Yeah. You have to, because you can get cancer if you get HPV virus…. It was helpful, because I know that now if I get the needle I won’t get cancer.”* Adolescent, ID8306, S10

Adolescents commonly expressed a need for more information about what to expect on vaccination day so that they could be prepared and manage their emotions. Adolescents often asked the researchers questions about the HPV vaccine, such as its purpose and benefits, why it was targeted at their age group only, how the vaccine works, why there was a need for second dose, what ingredients were in the vaccine, what the side effects could be, how long the vaccine would provide protection. These questions indicated that adolescents were curious and keen to understand the HPV vaccine. There was a common thread that the information provided to adolescents needed to go beyond that “*everyone just needs to get it*” (Adolescent, ID2819, S10); as one adolescent put it, “*there needs to be a reason*” (Adolescent, ID8306, S10).

Parents sometimes had similar information needs as adolescents regarding the basic facts about HPV vaccination, while others felt the information was adequate or that they did not need any further information. Some parents said they had signed up for the research because they wanted to find out more about the vaccination and that they and/or their adolescent had benefited from the chance to discuss issues about the vaccine through the research. They perceived the issue to be important because there was a research study being conducted about it. There were suggestions that similar opportunities to discuss the vaccine in a relaxed environment would benefit Aboriginal and Torres Strait Islander adolescents and their parents.

> *[Adolescent] loved the idea of having to be able to sit down with everyone and just be able to talk and stuff… So I reckon things like that would benefit the community more, because that way you know that you’re getting people actually in and they’re hearing you and us able to ask questions face-to-face…. Then that way the next person that goes and talks to someone else has got a more informed decision or more informed outlook, on what they’re spreading to other people when they’re talking about. It’s not misinformation being spread around.* Parent, ID5185, S10

Many were keen to empower their children with an understanding of the vaccination, suggesting skits, visual fact sheets, informal presentations and yarns as important educational tools.

> *You know, information is so powerful and them knowing that it’s to protect them and our mob, like kids, I think that would be very powerful within itself.* Parent, ID8400, S05

> *Especially for Indigenous kids, making sure that it’s very visual and that they can understand it. Not too many big health literacy words.* Parent, ID2452, S05

Some parents also suggested it would be beneficial to have a phone number to call for information from a trusted source about the vaccine. Convenience and immediacy of information was a priority, with some parents suggesting email or text messages as information and reminder systems, preferably from a trusted person at the school.

#### Parents led decision-making

For the vast majority of adolescents, parents made the final decision regarding whether they would receive the HPV vaccination. Parents generally felt it was appropriate that they make this decision. Adolescents trusted their parent’s decisions.

> *I don’t think, until they’re old enough to make their own decisions and they’re still under my roof, I will make the best decision I feel possible for my child, at that point in time. When they’re out on their own and they’re, you know, of age, then they can then make the decisions for themselves* Parent, ID3407, S03.

Adolescents shared that parents did not discuss the vaccination with them and many were not interested in having that discussion, despite expressing a need for more information about the vaccine. Boarding students had little to no opportunity to discuss whether they would receive the vaccine with their parents back home or at the residential college due to limited access to personal and communal phones. However, adolescents understood that parents made the decision for them to be vaccinated for their benefit.

> *Because we don’t really have a conversation about it. … My mum does everything to protect me from the sickness and that. She’s just, like, “You can get the needle.”* Adolescent, Yarning Circle 2, S02

#### HPV vaccination triggered “difficult conversations”

A handful of parents reported the HPV vaccinations triggering some serious conversations about sex, puberty, relationships, and cervical screening. Discussions about vaccinations were linked to adolescents maturing and learning more about the world, their bodies and relationships. One parent felt it was important to have these “difficult conversations” to ensure there was no shame associated with these topics.

> *It was a good chance to bring up around cervical cancer, Pap smears, all that sort of stuff. … Why women have [Pap smears] after they have had sex. … it’s taking that shame factor out as well…. That women’s health, trying to take that stigma away from it. [My child] hasn’t started her periods yet so it was a chance also to touch back up on that subject. So it was good. A good open up for us.* Parent, ID9932, S03

Others were aware of the potential for the vaccine to trigger these conversations and actively avoided the topic, instead emphasising the other health related benefits.

> *I’m trying to avoid having the sex talk with her at the moment, but I know that’s not far off. Yeah. But I just made her aware of how important it was to be able to have these needles and stuff like that. … I didn’t want to think the only reason she’s having that needle is just to be able to protect her, like safe sex kind of thing.* Parent, ID5185, S03

Both adolescents and parents felt that the school-based immunisation clinic was an opportunity for adolescents to demonstrate their emerging adulthood, particularly since the vaccination clinic was held in the first few months of adolescents’ first year of high school. Some adolescents were keen to demonstrate to parents and friends that they could manage the vaccination independently.

> *Well I said to her, “You’re going to have your needle today bub, do you want me to come?” She goes, “No, I’m at high school, I’m big now, I’ll be right.”* Parent, ID1439, S07.

### On vaccination day

While parents were most involved leading up to the day of the clinic, perhaps unsurprisingly adolescents held the strongest views about the day of the vaccination clinic.

#### Emotions ran high

For adolescents, emotions and feelings about the HPV vaccination centred on the experience of the vaccination clinic and administration of the vaccine. Most adolescents reported being scared and fearful of vaccinations, or at the very least not liking them. These feelings related to all vaccinations and needles rather than specifically the HPV vaccine and were driven by fear of, and prior experience of, physical pain from vaccinations. Adolescents with needle phobias were particularly distressed by the experience of the vaccine clinic at school. However, many adolescents were able to reconcile their worries about the vaccine with the health benefits.

Parents also reported their children being fearful of needles before the clinic but that many had reported afterwards that it had not been as bad as expected. Soreness at the vaccination site was the most common complaint. Many parents had discussed the short-term nature of the pain and reassured their child that it would pass quickly.

Many adolescents’ suggestions to improve HPV vaccination centred around distraction and relaxation while waiting for, receiving and recovering from the vaccination. For example, adolescents suggested putting a movie on, allowing students to use their phones for listening to music and playing games, calming music, colouring in, board games, and focusing on breathing. Others suggested reassurance and comforting techniques such as supporting their friends through the vaccination and ensuring that teachers, nurses and principals acknowledged that it may hurt and being responsive and empathetic to their feelings.

#### Need for privacy

Both adolescents and parents felt that worries about the vaccinations were heightened by the fact that vaccinations were administered in front of adolescents’ peers. The lack of privacy led to a high risk of embarrassment, shame, and teasing. Masses of adolescents waiting outside the room and/or being able to see the vaccinations being administered was a source of stress. In some cases where pinboards or whiteboards were placed as visual blockers, adolescents looked over or under them to view the clinic area. Many requested more privacy and protection from being seen by their friends and classmates to improve their comfort.

> *Because if it was in big groups like yesterday, we were all looking through the window and some of the kids started to panic because there was a big group watching. That’s how come I don’t like getting it done with groups of people, because if you start crying and that, they call you names, call you cry baby and that.* Adolescent, ID9022, S06

Some adolescents stated that they would prefer to have their vaccinations administered by their local doctor because it would provide a greater sense of privacy in the event they were upset by the vaccination. These participants generally would feel more comfortable with their doctor because they had known them a long time and trusted them.

Adolescents had many suggestions relating to improvements to privacy during the HPV vaccination. These included ensuring that the vaccination area was visually blocked from their classmates, avoiding big groups of people gathering for vaccinations and breaking classes down into small groups, administering needles one by one and spreading out the vaccination stations in the clinic.

#### Physical characteristics of the clinic

In addition to the importance of privacy, the room layout and clinic logistics impacted adolescents’ experience of vaccination. Poor clinic layout increased adolescents’ sense of anxiety and apprehension.

> *For me at the start, I was sort of like, it’ll be fine, it’s just a needle. But the chairs in the library, they were all lined up in alphabetical order, and I was last. So as everyone else was going, I was shuffling to the next chair and then looking in the door watching everyone, then the next chair.* Adolescent, Yarning Circle 3, S02

Observations of the clinics suggested that overlap in the waiting and recovery areas sometimes increased worries for those waiting: however, close supervision by a teacher or vaccination clinic staff helped to ameliorate this. In one school, some adolescents were connected by kinship and family ties to the vaccination clinic staff member overseeing a combined waiting and recovery area, which helped adolescents feel at ease and comfortable.

Schools employed a variety of strategies to keep adolescents calm and distracted from the vaccinations. For example, at one school the school chaplain supervised students in the recovery area and provided educational games for adolescents to play while seated. Another school played music throughout the entire clinic area, although this resulted in a loud vaccination clinic.

#### Trusted support people

The presence of trusted support people helped both adolescents and their parents to feel positive about the clinic, providing comfort, reassurance and advice. These included teachers, friends, vaccination clinic staff, and at times, parents.

At some schools, teachers running wellbeing and academic programs for Aboriginal and Torres Strait Islander adolescents were present at the vaccination clinic. In one case, an adolescent sought out one of these teachers to ask for support after a prior experience where the nurse was “rough” (Adolescent, ID8306, S10). Both the adolescent and parent reported that the adolescent had a more positive experience of the clinic because of support from the teacher.

Parents noted that the school-based vaccination clinic was challenging for some adolescents who were accustomed to having a parent or trusted person present to support them through the process, which caused distress.

> *She doesn’t like having sore arms or feeling queasy and I think it was a bit overwhelming for her this time, not having a parent as just a bit of a support system. She found that a bit overwhelming, but she got there in the end.* Parent, ID3407, S03

At one remote school, the presence of a family member and/or kinship connection was comforting to some children. This individual, known as Aka (Grandmother) to many adolescents at the school, was a staff member with the vaccination clinic and well known, respected and trusted in the community and this relationality was a comfort to adolescents.

### Q: How do you feel having Aka there to help you?

> *A: Good; supporting me through the day, and that stuff. … I feel safe. I feel comfortable that someone is there supporting me that I know.* Adolescent, ID7593, S09

Adolescents reported being reassured by the support of their peers and also providing support to others.

> *I told them to not [be] afraid of the needle because some friends are afraid of the needle. I told them not to fright, and when they get in they come out and they didn’t frighten, because I told them it’s [like a] mosquito bite.* Adolescent, ID5391, S09

Many adolescents suggested that the HPV vaccinations could be improved by having more support people available to help them.

Adolescents also reported negative experiences with teachers. For example, some adolescents talked about how a teacher had told them the needle would be big and teased them about how much it would hurt. This teacher also said that if they did not have the vaccinations, they would probably get sick. This caused more worries among adolescents.

#### Interactions with immunisers were brief and impersonal

Adolescents reported that the interactions with the nurse immuniser were fast and sometimes impersonal. While some could not remember what information was provided about the vaccines at the time, the overwhelming focus seemed to be on reassuring adolescents that the vaccination would be over quickly. Many adolescents reported being confused and embarrassed by the questions asked by the immuniser, especially regarding whether they were pregnant. There did not seem to be any context provided about this question and sometimes adolescents asked the research team why they had been asked this question.

Some adolescents would have preferred a more personable and educational approach.

> *I mean, I kind of would have liked to know a little bit more about it because people there, the ones that were giving us the needle, they had very small talk, they were like, “It’s only going to hurt for a bit,” and they didn’t really explain anything, they just said, “It’ll hurt like a little pinch and it’ll be over.” They didn’t really give any information about it.* Adolescent, ID2819, S10

## Discussion

The aim of this study was to understand the factors influencing the participation of Aboriginal and Torres Strait Islander adolescents in the school-based HPV vaccination program. A list of recommendations to support HPV vaccination among Aboriginal and Torres Strait Islander adolescents arising from the study findings is provided in Table 2.

**Table 2.**
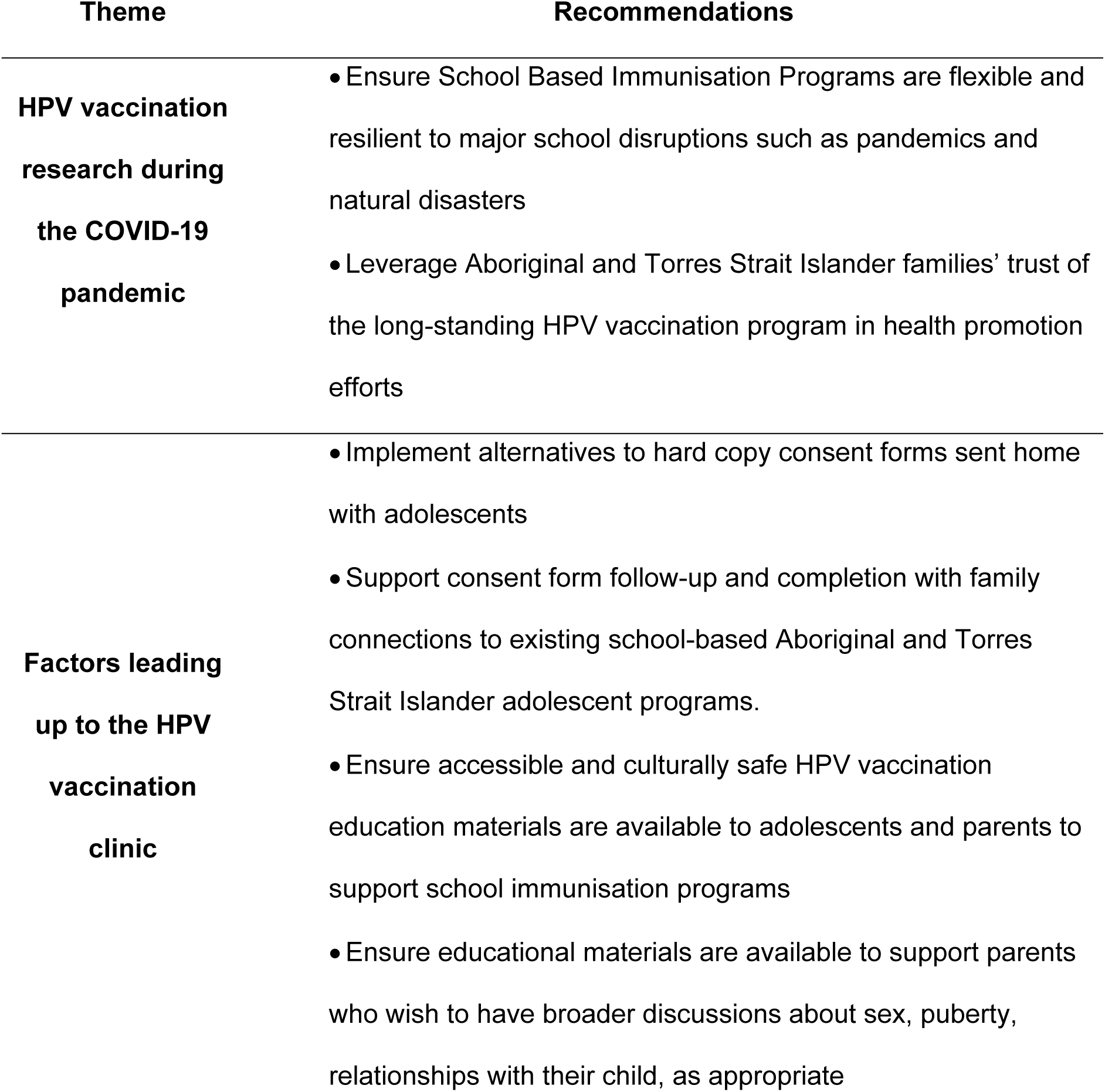
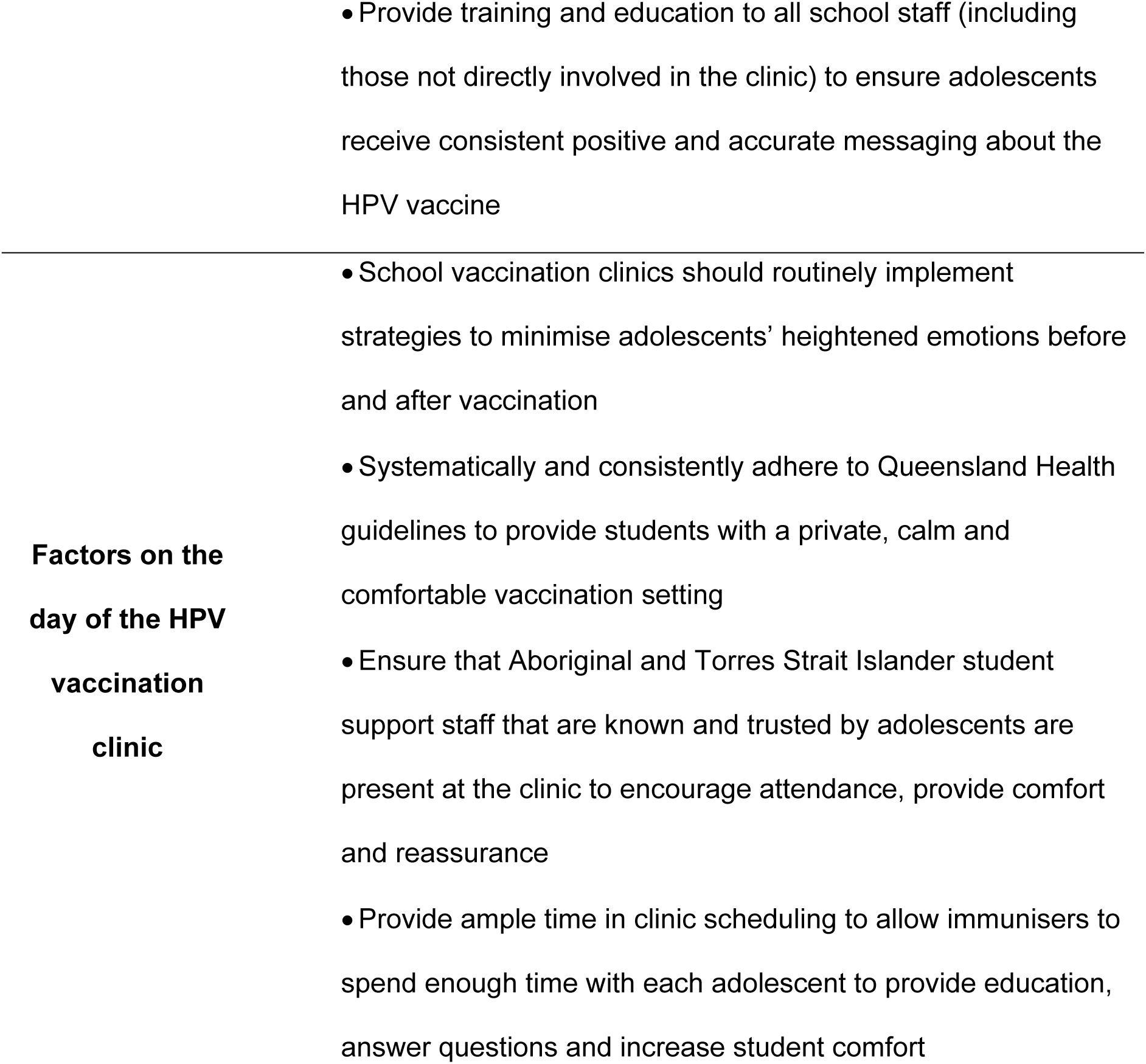
Recommendations to support HPV vaccination among Aboriginal and Torres Strait Islander adolescents.

This study was conducted during the COVID-19 pandemic and both parents and adolescents demonstrated heightened awareness of vaccination and significant support for vaccination for a range of reasons. Surprisingly, we found that participants felt the HPV vaccine was safer and more trustworthy than the COVID-19 vaccines, which were being developed and rolled out during the study period. This suggests that the long-standing nature of the HPV vaccination program and its anti-cancer messaging has been effective in building trust among Aboriginal and Torres Strait Islander communities. This contrasts with findings from other Indigenous populations where mistrust in government organisations is a key barrier to HPV vaccination uptake(13, 15). This is particularly important in the context of declining post-pandemic HPV vaccination uptake among both Aboriginal and Torres Strait Islander and non-Indigenous populations in Australia (3, 10) and globally(26), whereby recent gains in Aboriginal and Torres Strait Islander HPV vaccination coverage have been undone. Our findings suggest that declines in uptake may not be due to an increase in vaccine hesitancy and that with appropriate health promotion (discussed below), it may be possible to bring HPV vaccination uptake back up to pre-pandemic levels among Aboriginal and Torres Strait Islander communities. Notably school attendance amongst adolescents, has not returned to pre pandemic levels – for example school attendance (proportion of students with above 90% attendance) amongst Year 7 Aboriginal and Torres Strait adolescents in Queensland in 2023 was 79.8%, compared to 84.0% in 2019.

Similar to prior research in Australia and globally(27), adolescents and parents reported significant challenges in the consent process, with lost and forgotten forms common. Furthermore, there were calls for more accessible and culturally appropriate health promotion materials for both parents and adolescents, delivered in engaging ways. An audit of HPV vaccination materials for Aboriginal and Torres Strait Islander audiences found a handful of resources but it was unclear how, if it at all, they were being used to support school immunisation clinics(28). Our findings indicate there is a need for place-based methods to support Aboriginal and Torres Strait Islander families to complete and return the consent form; this may include digital or telephone consent options, supported by Aboriginal and Torres Strait Islander student support staff or organisations, consistent with prior research.(29) There are also opportunities for development of short, accessible HPV vaccination information sheets, videos, or other modes of information sharing, embedded within the delivery of the school immunisation program, to support Aboriginal and Torres Strait Islander communities to make informed decisions about HPV vaccination. These resources must recognise the collective and relational perspectives of Aboriginal and Torres Strait Islander communities in terms of how information is evaluated and decisions made. Critically, funding must be dedicated to maintain and sustain these materials over time to ensure ongoing accuracy and support for this important vaccination program. Furthermore, the findings suggest there was a need for all school staff, including those not directly involved in the clinic, to complete a standard package of education and expectations on their support role to ensure adolescents and parents receive consistent, accurate and positive messaging about vaccination clinics.

This study found that a lack of privacy and poor clinic layout negatively impacted Aboriginal and Torres Strait Islander adolescents’ experience of the vaccination clinic. It is critical that the vaccination is private, and the clinic is set up to avoid potential shame and embarrassment triggered by feeling observed by peers. In Queensland, the set-up of immunisation clinics is dictated by rigorous protocols to ensure the safe, effective and efficient delivery of vaccines. Schools receive an information booklet(30) that outlines clinic providers’ requirements for the clinic set up. Our findings suggest that, despite best efforts under time-pressured conditions, clinic layout guidelines are inconsistently adhered to. School staff and immunisation providers also reported challenges in the clinic set up and delivery in the school setting (31). Schools and immunisation providers may require more support and monitoring to ensure clinics do not cause adverse effects triggered by the process of vaccination as seen in other Australian settings.(32) Effective clinic set up can support a culturally safe and positive vaccination experience for Aboriginal and Torres Strait Islander adolescents.

We found that the presence of trusted support people helped Aboriginal and Torres Strait Islander adolescents to have a more positive experience of the HPV vaccination. These included a range of people such as Aboriginal and Torres Strait Islander student support staff, teachers, friends and parents. Systematic integration of dedicated support staff into the delivery of the clinic may improve the experience of vaccination for these adolescents. Furthermore, involving Aboriginal and Torres Strait Islander support staff and organisations in the facilitation of information and collection of consent forms may also improve community engagement and ultimately uptake of the HPV vaccine.

This research was conducted while the two-dose schedule of the HPV vaccine was current. Since January 2023, HPV vaccination has been delivered with a single dose. We found that many people were unaware of the need for a second dose and that declining school attendance at the end of the school year may prevent high 2^nd^ dose coverage. Therefore, the new one-dose schedule is likely to facilitate equity in HPV vaccination coverage among Aboriginal and Torres Strait Islander communities as long as opportunities for vaccination are maintained, including catch-up clinics when needed.

### Strengths and limitations

This project is the only study of Aboriginal and Torres Strait Islander adolescents’ and parents’ views on school-based HPV vaccination. It was strengthened by an Indigenist research approach complemented by structuring the analysis around the socio-ecological model for health promotion. The study was conceptualised, conducted, and analysed with the leadership of Aboriginal and Torres Strait Islander peoples. This is important given few studies reporting on the views of Aboriginal and Torres Strait Islander adolescents and parents, and the lack of research led by Indigenous communities globally.(15) This study had limited participation from unvaccinated adolescents and their parents.

Therefore the recommendations provided in Table 2 may not include strategies to support unvaccinated adolescents and their parents. While important insights have been gained from this largely vaccinated cohort, it has missed insights from unvaccinated adolescents and parents, which may be useful to enhance the program. Due to school and clinic adaptations due to the pandemic, the findings may not represent “business as usual”: however, they provide insight into how Aboriginal and Torres Strait Islander families may approach HPV vaccination in the context of heightened awareness of vaccination. Finally, self-reported HPV vaccination status (Table 1) is difficult to interpret as it was influenced by the timing of the fieldwork and we did not verify vaccination status.

### Conclusions

Globally, Indigenous peoples carry a high burden of HPV infection.(33) HPV vaccination through school programs has great potential to minimise this burden on Aboriginal and Torres Strait Islander peoples, with coordinated and sustained action now required if we are to reach the target of 90% vaccination set by the National Strategy for the Elimination of Cervical Cancer in Australia.(3) This is particularly important in light of declining uptake of HPV vaccination in many countries in a post-COVID-19 world. The findings of this study provide clear recommendations for policy and practice to improve HPV vaccination among Aboriginal and Torres Strait Islander peoples and contribute to equity in HPV-related cancer outcomes.

## Data Availability

The datasets generated and/or analysed during the current study are not publicly available due to ethical and confidentiality reasons. Participants did not consent to public release of full transcripts and the nature of the qualitative data are such that public availability would compromise participant confidentiality. Requests for specific excerpts relating to published verbatim quotes will be considered on a case-by-case basis. Requests for data may be sent to the corresponding author and to the Human Research Ethics Committee (HREC) of the Northern Territory Department of Health and Menzies School of Health Research ( reference number: 2019-3484). Please note that the authors will need to seek approval from the six HRECs listed in the Ethics Approval section before releasing data.

## Acknowledgements

We wish to thank Royden Fagan (RF), Evan AhWing (EA) and Elsie Seriat (ES) for their assistance in data collection.

We also acknowledge the guidance, feedback and advice from the Aboriginal and Torres Strait Islander Steering Committee: Vanessa Clements, Evan AhWing, Frances Lomas, Casey Ross, and Sonya Egert.

Informants retain ownership of Aboriginal and Torres Strait Islander knowledge and cultural heritage.

## Supporting information

**S1 File. COREQ checklist.**

**S2 File. CONSIDER checklist**

**S3 File. School Characteristics**

**S4 File. Yarning Guides**

## References

1. Hall MT, Simms KT, Lew J-B, Smith MA, Brotherton JML, Saville M, et al. The projected timeframe until cervical cancer elimination in Australia: a modelling study. Lancet Public Health. 2019;4(1):e19–e27.

2. Australian Bureau of Statistics. Estimates and Projections, Aboriginal and Torres Strait Islander Australians Canberra: ABS; 2024 [Available from: https://www.abs.gov.au/statistics/people/aboriginal-and-torres-strait-islander-peoples/estimates-and-projections-aboriginal-and-torres-strait-islander-australians/latest-release.

3. Machalek D, Smith M, Brotherton J, Canfell K, Pagotto A, Saville AM, et al. Cervical Cancer Elimination Progress Report: Australia’s progress towards the elimination of cervical cancer as a public health problem. 2023 09/05/2024.

4. Cancer Council. About HPV and cancer - HPV vaccine 2023 [Available from: https://www.hpvvaccine.org.au/hpv-and-cancer/about-hpv-and-cancer.

5. Lei J, Ploner A, Elfstrom KM, Wang J, Roth A, Fang F, et al. HPV Vaccination and the Risk of Invasive Cervical Cancer. N Engl J Med. 2020;383(14):1340–8.

6. Australian Government Department of Health and Aged Care. National Immunisation Program Schedule Canberra: Australian Government; 2024 [Available from: https://www.health.gov.au/topics/immunisation/when-to-get-vaccinated/national-immunisation-program-schedule.

7. National Centre for Immunisation Research & Surveillance. Significant events in human papillomavirus (HPV) vaccination practice in Australia 2023. 2023.

8. Australian Government Department of Health and Aged Care. Human papillomavirus (HPV) vaccine: Changes under the National Immunisation Program 2023 [

9. Australian Centre for the Prevention of Cervical Cancer. National Strategy for the Elimination of Cervical Cancer in Australia. 2023.

10. National Centre for Immunisation Research & Surveillance Australia. Annual Immunisation Coverage Report 2023. Westmead: NCIRS; 2023.

11. Vujovich-Dunn C, Wand H, Brotherton JML, Gidding H, Sisnowski J, Lorch R, et al. Measuring school level attributable risk to support school-based HPV vaccination programs. BMC Public Health. 2022;22(1):822.

12. Lockwood L, Ju X, Sethi S, Hedges J, Jamieson L. Knowledge and Awareness of HPV, the HPV Vaccine and Cancer-Related HPV Types among Indigenous Australians. Int J Environ Res Public Health. 2024;21(3).

13. Poirier B, Sethi S, Garvey G, Hedges J, Canfell K, Smith M, et al. HPV vaccine: uptake and understanding among global Indigenous communities - a qualitative systematic review. BMC Public Health. 2021;21(1):2062.

14. Whop LJ, Smith MA, Butler TL, Adcock A, Bartholomew K, Goodman MT, et al. Achieving cervical cancer elimination among Indigenous women. Prev Med. 2021;144:106314.

15. MacDonald SE, Kenzie L, Letendre A, Bill L, Shea-Budgell M, Henderson R, et al. Barriers and supports for uptake of human papillomavirus vaccination in Indigenous people globally: A systematic review. PLOS Glob Public Health. 2023;3(1):e0001406.

16. Whop LJ, Butler TL, Brotherton JML, Anderson K, Cunningham J, Tong A, et al. Study protocol: Yarning about HPV Vaccination: a qualitative study of factors influencing HPV vaccination among Aboriginal and Torres Strait Islander adolescents in Australia. BMJ Open. 2021;11(8):e047890.

17. Rigney L. Internationalization of an Indigenous Anticolonial Cultural Critique of Research Methodologies: A Guide to Indigenist Research Methodology and Its Principles. Wicazo Sa Review. 1999;14(2).

18. Rigney L. Indigenous Australian views on Knowledge production and Indigenist research. In: Kunnie J, Goduka I, editors. Indigenous Peoples’ Wisdom and Power: Affirming Our Knowledge Through Narratives. Burlington: Ashgate; 2006. p. 32-49.

19. McLeroy KR, Bibeau D, Steckler A, Glanz K. An ecological perspective on health promotion programs. Health Educ Behav. 1988;15(4):351–77.

20. Tong A, Sainsbury P, Craig J. Consolidated criteria for reporting qualitative research (COREQ): a 32-item checklist for interviews and focus group. Int J Qual Health Care. 2007;19:349–57.

21. Huria T, Palmer SC, Pitama S, Beckert L, Lacey C, Ewen S, et al. Consolidated criteria for strengthening reporting of health research involving indigenous peoples: the CONSIDER statement. BMC Med Res Methodol. 2019;19(1):173.

22. Australian Curriculum AaRA. ACARA 2024 [Available from: https://www.acara.edu.au/.

23. Bessarab D, Ng’andu B. Yarning About Yarning as a Legitimate Method in Indigenous Research. International Journal of Critical Indigenous Studies. 2010;3(1).

24. Braun V, Clarke V. Using thematic analysis in psychology. Qual Res in Psychol. 2006;3(2):77–101.

25. Queensland Health. School Immunisation Program: Queensland Health; 2024 [Available from: https://www.health.qld.gov.au/clinical-practice/guidelines-procedures/diseases-infection/immunisation/schools.

26. Casey RM, Akaba H, Hyde TB, Bloem P. Covid-19 pandemic and equity of global human papillomavirus vaccination: descriptive study of World Health Organization-Unicef vaccination coverage estimates. BMJ Med. 2024;3(1):e000726.

27. Ferrer HB, Trotter C, Hickman M, Audrey S. Barriers and facilitators to HPV vaccination of young women in high-income countries: a qualitative systematic review and evidence synthesis. BMC Public Health. 2014;14.

28. Butler TL, Morseu-Diop A, Brotherton JML, Peart L, Jayasekara I, Peart A, et al. An evaluation of Human Papillomavirus vaccination resources available to Aboriginal and Torres Strait Islander adolescents and parents and caregivers in Australia [Manuscript submitted for publication]. 2024.

29. Swift C, Dey A, Rashid H, Clark K, Manocha R, Brotherton J, et al. Stakeholder Perspectives of Australia’s National HPV Vaccination Program. Vaccines (Basel). 2022;10(11).

30. Queensland Health. Queensland School Immunisation Program—Information for Schools. State of Queensland: Queensland Health; 2023.

31. Morseu-Diop A, Butler TL, Anderson K, Brotherton JML, Cunningham J, Jaure A, et al. Stakeholder perspectives on HPV vaccination uptake among Aboriginal and Torres Strait Islander adolescents via the School Immunisation Program in Queensland [Manuscript submitted for publication]. 2024.

32. Buttery JP, Madin S, Crawford NW, Elia S, La Vincente S, Hanieh S, et al. Mass psychogenic response to human papillomavirus vaccination. Med J Aust. 2008;189(5):261–2.

33. Sethi S, Ali A, Ju X, Antonsson A, Logan R, Canfell K, et al. A systematic review and meta-analysis of the prevalence of human papillomavirus infection in Indigenous populations - A Global Picture. J Oral Pathol Med. 2021;50(9):843–54.

